# Probiotic Supplementation Improves Short-Chain Fatty Acids Production in Malnourished Children: A Systematic Review and Meta-Analysis

**DOI:** 10.1101/2025.06.22.25330094

**Authors:** Rizqi Yanuar Pauzi, Laksita Widya Kumaratih, Puspita Tri Yuliana, Raghda Aisy Aqila

**Affiliations:** Department of Microbiology, Faculty of Medicine, Universitas Jenderal Soedirman, Purwokerto 53147, Indonesia; Faculty of Medicine, Universitas Jenderal Soedirman, Purwokerto 53147, Indonesia

**Keywords:** probiotic, short-chain fatty acids, malnutrition, stunting, children, meta-analysis

## Abstract

This systematic review and meta-analysis aimed to asses the impact of probiotic supplementation on short-chain fatty acids (SCFAs) production such as acetate, propionate, and butyrate in malnourished children. We hypothesize that probiotic supplementation would significanty improve SCFAs production in malnourished children. We conducted a comprehensive search in Scopus, PubMed, Google Scholar, Semantic Scholar, and Crossref up to February 2025. We included randomized controlled trials (RCTs) that involved children with undernutrition receiving probiotic supplementation. Risk of bias was assessed using the Cocrane risk-of-bias tool (RoB 2.0) and certainty of evidence was evaluated using the grading of recommendations assessment, development, and evaluation (GRADE) approach. Meta-analyses and meta-regressions were performed using R statistical software. A total of three RCTs involving 378 children were included in the analysis. Probiotic supplementation may improve overall SCFAs production in malnourished children, although the effect was not statistically significant for total SCFAs or for individual SCFAs such as acetate and propionate. However, a significant increase in butyrate levels was observed following probiotic supplementation (MD = 0.63, 95% CI: 0.14–1.12). Subgroup analyses showed no significant differences across SCFA types, indicating consistent effects of probiotics on SCFA profiles. Meta-regression analyses revealed that factors such as sample size, intervention duration, age, and dosage did not significantly moderate the outcomes. These findings suggest that probiotics may serve as a potential nutritional strategy for improving gut microbiota function in malnourished children.

## Introduction

Malnutrition is a major public health concern, especially in developing countries like Indonesia. It is broadly divided into two categories: overnutrition and undernutrition. Children’s undernutrition is commonly classified into three forms based on world health organization growth standards: stunting (low height-for-age), wasting (low weight-for-height), and underweight (Gunawan et al. 2021). Stunting indicates chronic malnutrition, wasting reflects acute malnutrition, and being underweight may be a combination of the two (Thurstans et al. 2022). We use Z-score thresholds to assess severity. Moderate malnutrition is defined as values between -2 and -3 standard deviations (SD), and severe malnutrition as values less than -3 SD (Manary and Sandige 2008). In Indonesia, the prevalence of stunting among children under five has risen to 21.5%, with wasting at 7.7% and underweight at 17.1% (Kemenkes 2022).

The development of undernutrition is influenced by a range of interrelated factors, including inadequate dietary intake, frequent infections, poor feeding practices, and limited access to sanitation and clean water (Shrestha et al. 2020). These underlying causes not only compromise physical growth but also impair cognitive development and immune system function and increase vulnerability to disease and mortality (Black et al. 2013). Furthermore, emerging evidence highlights the role of gut microbiota in the development and outcomes of undernutrition (Blanton et al. 2016). Alterations in the composition and function of gut microbes can impair nutrient metabolism and immune responses (Wolowczuk et al. 2008). The gut microbiota breaks down dietary fibers into SCFAs, especially acetate, propionate, and butyrate, which are important for keeping the gut healthy, supporting the immune system, and promoting overall growth and development (Subramanian et al. 2014).

SCFAs, mainly acetate, propionate, and butyrate, are the main products made by microbes in the colon and are connected to different body functions (Fusco et al. 2023). Acetate plays a role in lipid metabolism and immune regulation, propionate contributes to gluconeogenesis and satiety signaling, while butyrate serves as the main energy source for colonocytes and exhibits strong anti-inflammatory effects (Yoshida, Ishii, and Akagawa 2019; Jiao et al. 2021; Salvi and Cowles 2021; Liu, Fu, and Li 2019). The gut bacteria in stunted children have been found to produce less SCFAs, which may result in poor nutrient absorption, problems with the gut barrier, and a higher risk of gut infections (Surono et al. 2024). Based on these findings, efforts to restore the balance of gut bacteria and boost SCFAs production are especially important for tackling the health issues caused by undernutrition.

Probiotics are live microorganisms that have beneficial impacts on health when consumed in adequate amounts (Kamil et al. 2022). There are several types of probiotics, including species such as *Lactobacillus*, *Bifidobacteria*, and *Saccharomyces* (Famouri et al. 2014). The consumption of probiotics can help maintain the balance of gut microbiota, enhance the body’s immune system, and provide various other health benefits (Rahayu et al. 2024). Probiotics can increase the number of beneficial bacteria and reduce the number of less beneficial or pathogenic bacteria by increasing the production of SCFAs, which play a role in lowering the pH levels of the intestinal environment and thereby preventing the colonization of pathogenic bacteria. Additionally, SCFAs also help improve nutrient absorption in the intestines (Gunawan et al. 2021; Markowiak-Kopeć and Śliżewska 2020). Several previous studies have shown that probiotic supplementation can increase body weight, height, and body mass index, indicating that probiotic administration can support growth in undernourished children (Gunawan et al. 2021; Borgeraas et al. 2018).

This systematic review and meta-analysis evaluates the impact of probiotic supplementation on SCFAs levels in stunted children. It aims to determine efficacy compared to controls, explore sources of heterogeneity, and assess evidence quality. We hypothesize that probiotics significantly increase SCFAs production. This review offers a comprehensive synthesis to inform the role of probiotics as a nutritional strategy for malnourished children.

## Methods

This systematic review and meta-analysis adheres to the protocols established in the *Cochrane Handbook for Systematic Reviews of Interventions* (Higgins et al. 2019) and adheres to the reporting criteria delineated by the *Preferred Reporting Items for Systematic Reviews and Meta-Analyses* (PRISMA) (Moher et al. 2016). The study protocol was registered with the *International Prospective Register of Systematic Reviews* (PROSPERO) on April 22, 2025, under registration code CRD420251036758, before the study commenced.

### Eligibility Criteria

Studies were chosen according to predetermined inclusion and exclusion criteria. Eligible studies must include children aged 0 to 12 years who have been diagnosed with undernutrition, which is defined by world health organization growth standards as having one or more of the following: stunting (height-for-age Z-score below -2 SD), wasting (weight-for-height Z-score below -2 SD), or being underweight. The intervention of interest was supplementation with probiotics, considering factors such as dosage, type of supplementation, length of usage, and dosing intervals, whether delivered alone or in combination. The control group may consist of any similar cohort, including no-intervention, placebo, or alternative pharmacological supplement. The primary outcomes of interest included acetate, propionate, and butyrate levels, which were assessed in feces or serum. Only RCTs published in peer-reviewed journals were included. The following studies were excluded: (1) observational studies (e.g., cohort, case-control, or cross-sectional designs); (2) studies with incomplete or missing SCFAs data; and (3) animal studies or in vitro experiments.

### Search Strategy

We conducted a comprehensive literature search across five major electronic databases: Scopus, PubMed, Google Scholar, Semantic Scholar, and Crossref. The search included studies published from inception to February 2025. We used the following Medical Subject Headings (MeSH) terms and keywords to identify relevant studies: ("Probiotics" OR "Lactobacillus" OR "Bifidobacterium" OR "Saccharomyces" OR "Streptococcus" OR "Probiotics [MeSH Terms]" AND ("Short-Chain Fatty Acids" OR "SCFA" OR "acetate" OR "propionate" OR "butyrate" OR "Short-Chain Fatty Acids [MeSH Terms]") AND ("Stunting" OR "Growth Retardation" OR "Malnutrition" OR "Undernutrition" OR "Failure to Thrive" OR "Growth Failure" OR "Stunting [MeSH Terms]"). Additionally, reference lists of relevant systematic reviews and meta-analyses were manually searched to identify additional eligible studies. No language restrictions were applied during the initial search phase.

### Study Selection Process

All identified studies were imported into Rayyan, a web-based platform to facilitate the screening process. All identified studies were independently screened by three reviewers (LWK, PTY, and RAA) based on titles and abstracts. Full-text screenings were conducted for studies that met the initial eligibility criteria. Discrepancies between reviewers were resolved by a fourth reviewer (RYP). The study selection process was documented using a PRISMA flow diagram, detailing the number of studies included and excluded at each stage of screening.

### Data Extraction and Risk of Bias Assessment

Data were extracted in a uniform format to ensure consistency and accuracy when synthesizing the findings. The retrieved data contained crucial study features such as the author’s name, year of publication, nation of study, and study methodology used. Participants’ details were collected, including their ages, sample size, and the study’s precise definition of stunting. Details about the intervention were documented, including the type of probiotic strain used, the dosage, and the duration of administration. The key outcome measures of interest were SCFAs levels in fecal or serum samples, which included acetate, propionate, and butyrate. Furthermore, the retrieved data includes information such as mean differences in SCFAs levels between intervention and control groups, as well as standard deviations and effect sizes to aid in meta-analysis.

The potential for bias in all included studies was independently evaluated utilizing the Cochrane Handbook’s Risk of Bias Version 2 (RoB 2.0) tool (Sterne et al. 2019). For the risk of bias assessment, two writers (PTY and RAA) served as primary reviewers, with LWK as secondary reviewer. Disputes were settled by dialogue or by consulting a third reviewer (RYP). The RoB 2.0 tool assesses five domains: randomization bias, variations from intended interventions, missing outcome data, assessment of outcomes, and selection of the reported result. We rated the risk of bias in each study as low, with some concerns, or high. We used the GRADE method to determine the certainty of evidence. This evaluation was performed autonomously by two authors (LWK and RAA), with PTY assisting in the resolution of any disputes. GRADE takes into account five factors: bias, inconsistency, indirectness, imprecision, and publication bias. The quality of the evidence was classified as excellent, moderate, low, or very low (Oxman 2004).

### Data Synthesis and Statistical Analysis

A comprehensive meta-analysis was performed using R statistical software (v.4.5.0) (R Core Team 2022) to integrate the quantitative findings from the included studies. The analysis aimed to assess the impact of probiotic supplementation on SCFAs production in malnourished children. For continuous outcomes, such as the specific SCFAs levels (acetate, propionate, and butyrate), mean differences (MD) with 95% CIs were calculated.

Subgroup analyses were performed to investigate the possible differences in the outcomes of SCFAs level depending on the studies’ major features. This involved the comparison of several outcomes, including acetate, propionate, and butyrate. The I² statistic was employed to evaluate heterogeneity among research, indicating the variety in outcomes among them. When I² values above 50% and the p-value from the chi-squared test was below 0.05, signifying substantial heterogeneity, a random-effects model was employed in lieu of a fixed-effects model to accommodate the variability. This methodology enabled a more adaptable analysis, accounting for variations in research design, demographics, and treatments.

Additionally, a meta-regression was conducted using metafor R package (v.4.8-0) (Viechtbauer 2010) to explore potential factors that could explain the observed heterogeneity. This regression examined variables such as total sample size, duration of intervention, participant age, and dosage of intervention. This analysis aimed to identify which study characteristics most significantly influenced the outcomes.

The meta-analysis concentrated on the synthesis of SCFAs such as acetate, propionate, and butyrate levels. We assembled a summary of the data in tabular style to present an orderly overview of the individual research outcomes. This table facilitated a more lucid comparison of the studies’ results and underscored any patterns or discrepancies in the impact of various treatments. This thorough methodology facilitated an in-depth comprehension of the potential effects of probiotics short-chain fatty acid synthesis in malnourished children, emphasizing the quality and reliability of the existing data..

## Results

### Study Selection

The systematic review began with a comprehensive search across multiple databases and registers, yielding a total of 4,436 records. These records were sourced from Scopus (n=141), PubMed (n=40), Google Scholar (n=3,198), Semantic Scholar (n=57), and Crossref (n=1,000). After removing 1,647 duplicate records, 100 records flagged as ineligible by automation tools, and 10 records excluded for other reasons, 2,679 records remained for screening. During the title and abstract screening phase, 2,665 records were excluded as they did not meet the inclusion criteria. This phase left 14 full-text articles for detailed eligibility assessment. Of these, 11 articles were excluded for the following reasons: 8 did not assess relevant outcomes, 2 focused on healthy children rather than the target population, and 1 had incompatible study designs. Ultimately, 3 studies met all eligibility criteria and were incorporated into the systematic review and meta-analysis, as illustrated in Fig. 1.

**Fig. 1.**
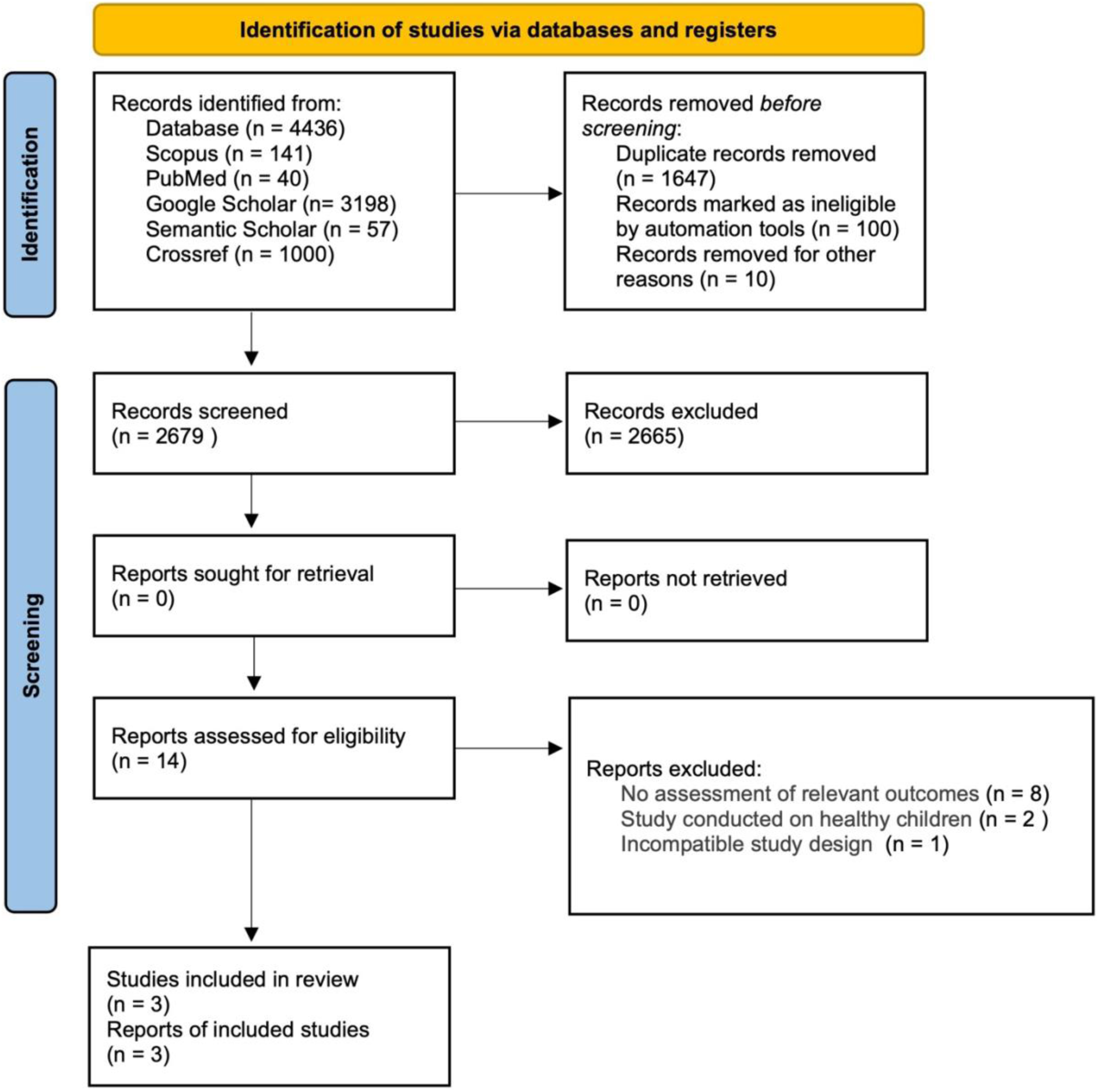
PRISMA flow diagram illustrating the study selection process. The flowchart outlines the step-by-step process of identifying, screening, and selecting eligible randomized controlled trials (RCTs) evaluating the effects of probiotic interventions on short-chain fatty acid (SCFA) production in malnourished children. A total of 3 studies were included after removing duplicates and applying inclusion/exclusion criteria. RCT, randomized controlled trial; SCFA, short-chain fatty acid.

### Characteristics of Included Studies

The systematic review included three randomized controlled trials published between 2021 and 2024, conducted in Indonesia (Kamil et al. 2022; Rahayu et al. 2024; Rahayu et al. 2021). These studies involved a total of 378 participants, with sample sizes ranging from 30 to 56 children. The age of participants varied widely, from infants to 12 years as shown in Table 1. The probiotic strains studied were *Lactobacillus plantarum* Dad-13. Delivery methods included powders (1 study) and gummies/chocolates (2 studies). Intervention durations spanned from 50 to 90 days. Key outcomes assessed were improvements in acetate, propionate, and butyrate.

**Table 1.** Baseline characteristics of included studies investigating the impact of probiotic supplementation on SCFAs levels in malnourished children.

| Author, year, nation | Research Methodology | Study population | Participants' age | Intervention | Control | Intervention Duration | Results |
| --- | --- | --- | --- | --- | --- | --- | --- |
| Kamil et al., (2022)<br>Indonesia | Randomized double-blind controlled trial | Probiotic (n=15)<br>Placebo (n=15) | Infant | Gummy made from skim milk powder containing *L. plantarum* Dad-13 ($8.96 \times 10^8$ - $1.16 \times 10^9$ CFU/3 g)/day | Gummy made from skim milk powder (Lactona) | 50 days | Both placebo and probiotic groups showed a significant increment in body weight and height. In addition, elevation of total SCFAs. |
| Rahayu et al., (2024)<br>Indonesia | Randomized, double-blinded, placebo-controlled trial | Probiotic (n=28)<br>Placebo (n=28) | 8-12 years | A probiotic chocolate containing chocolate and an indigenous probiotic, *L. plantarum* Dad-13 ($1 \times 10^9$ CFU/bar), which is consumed once a day | Chocolate with the same composition without probiotics | 90 days | It does not significantly affect body weight, height, or SCFAs. |
| Rahayu et al., (2021)<br>Indonesia | Randomized, double-blind, placebo-controlled design | Probiotic (n=20)<br>Placebo (n=20) | 10-12 years | Probiotic powder containing *L. plantarum* Dad-13 of $1 \times 10^9$ CFU/bar) 1 gram/day | Skim milk powder from a commercial product | 60 days | Synbiotics may increase the weight, height, and SCFAs of children. |

### Risk of Bias Assesment

The primary sources of bias identified across the included studies were randomization process. Among the three studies, two were assessed as having a low risk of bias, although one exhibited some concerns; none were classified as having a high risk of bias as shown in Fig. 2 and 3.

**Fig. 2.**
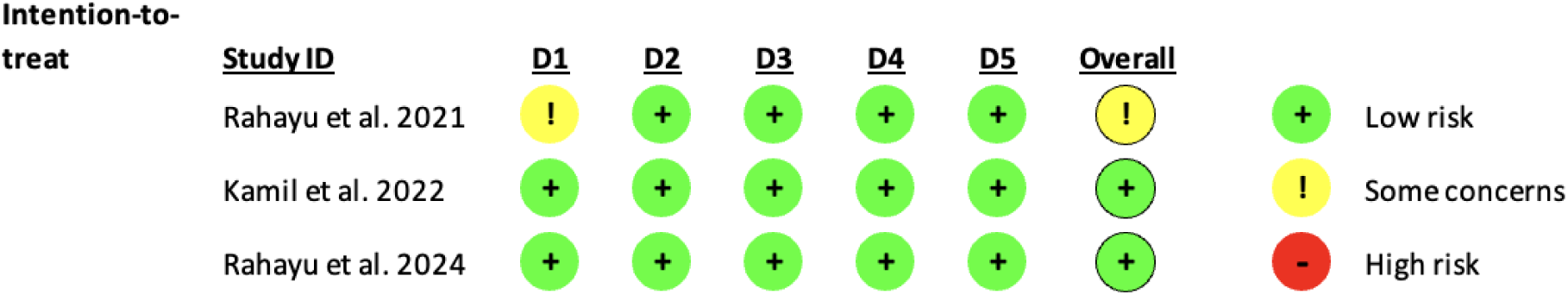
Risk of bias assessment across studies using RoB 2.0 tool. The overall risk of bias among the three included studies was assessed based on five domains: randomization process (D1), deviations from intended interventions (D2), missing outcome data (D3), measurement of the outcome (D4), and selection of the reported result (D5). Two studies showed low risk, while one study classified as some concerns.

**Fig. 3.**
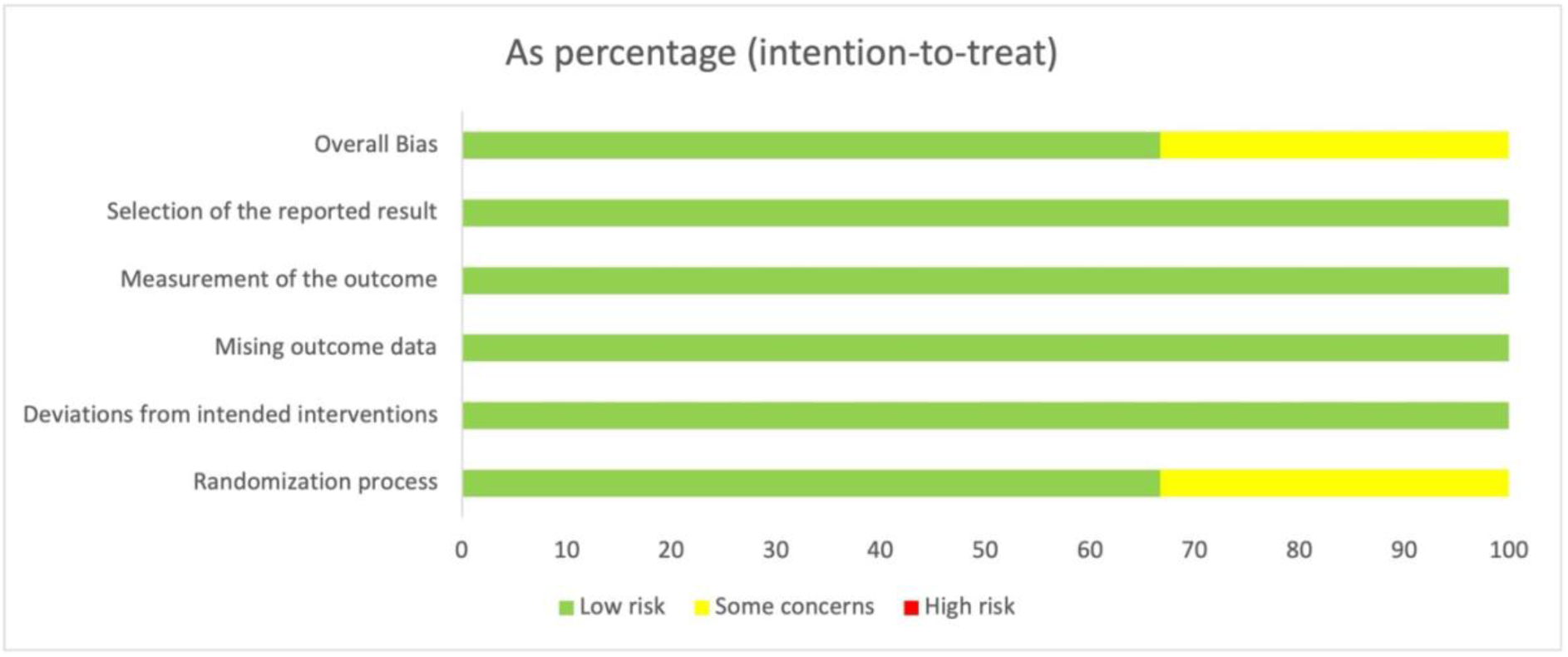
Distribution of risk of bias assessment results. Percentage distribution of low risk, some concerns, and high risk across each domain of bias assessed in the included studies.

### Meta-analysis

#### Overall Effect on SCFAs

Probiotic therapies in malnourished children may elevate total SCFA levels relative to the control group. The meta-analysis yielded a mean difference (MD) of 0.40 (95% CI: -0.12 to 0.93), based on three trials involving 378 children. The heterogeneity across studies was low (I² = 11.7% ), indicating consistency in the findings. Subgroup analysis further revealed that the outcomes for individual SCFAs (acetate, propionate, and butyrate) did not exhibit significant differences across subgroups (χ² = 5.53, df = 2, p = 0.0629 ) as seen in Fig. 4. This suggests that the overall effect of probiotics on SCFAs is consistent across these components.

**Fig. 4.**
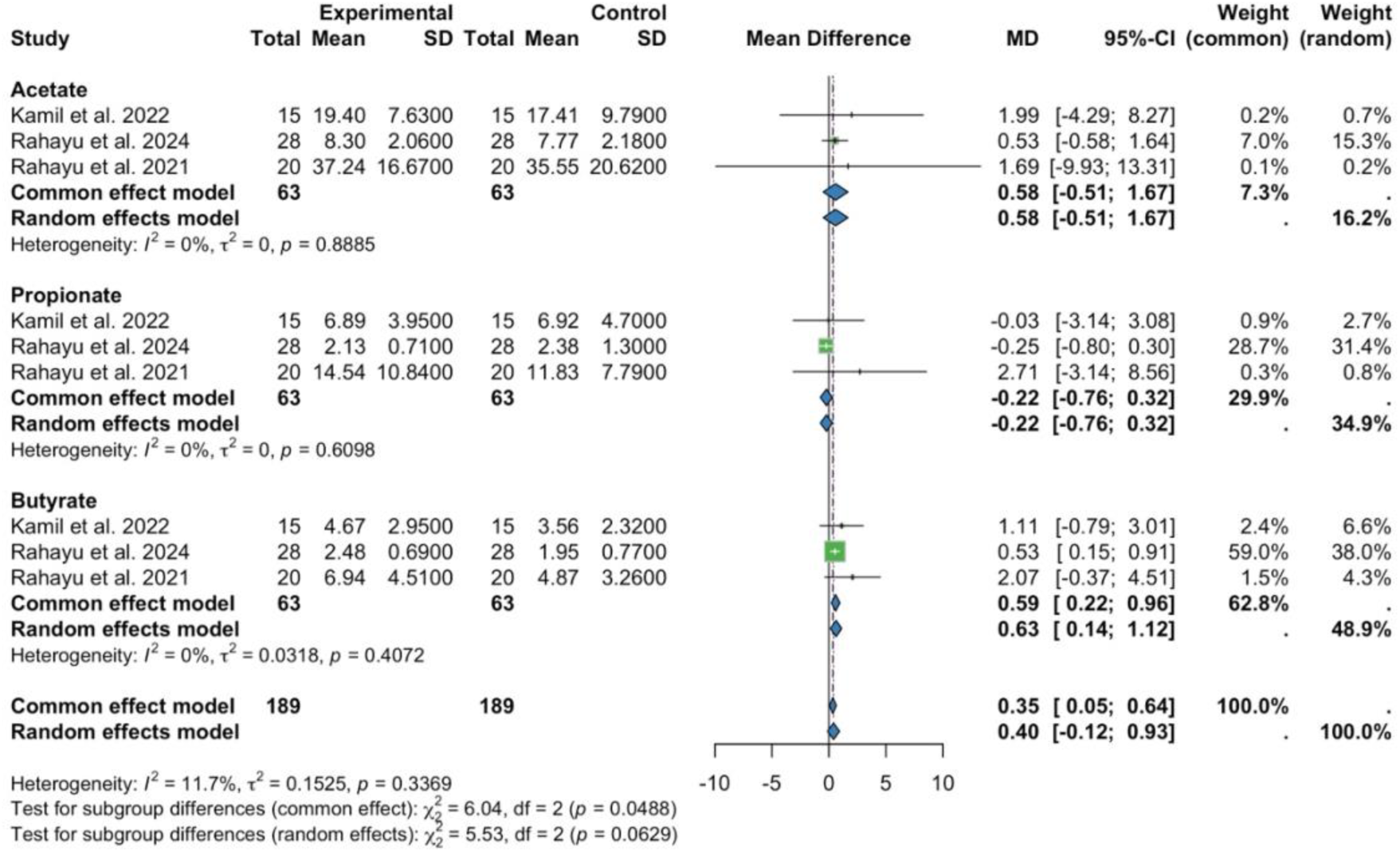
Impact of probiotic supplements on SCFAs levels. Forest plot showing the mean differences (MD) and 95% confidence intervals (CI) for acetate, propionate, and butyrate levels between experimental and control groups across three studies.

#### Acetate

Probiotic therapies appear to have a modest effect on elevating acetate levels in malnourished children. The meta-analysis reported a mean difference (MD) of 0.58 (95% CI: -0.51 to 1.67) based on three trials involving 126 children. However, the confidence interval includes zero, indicating that this effect is not statistically significant. There was no heterogeneity among studies (I² = 0% ). Meta-regression analysis indicated that total sample size (p = 0.6278), length of the intervention (p = 0.6265), age (p = 0.6629), and dose of intervention (p = 0.6577) were not significant moderators for acetate levels, as presented in Table 2. This suggests that the observed effect of probiotics on acetate levels is not influenced by these variables.

**Table 2.**
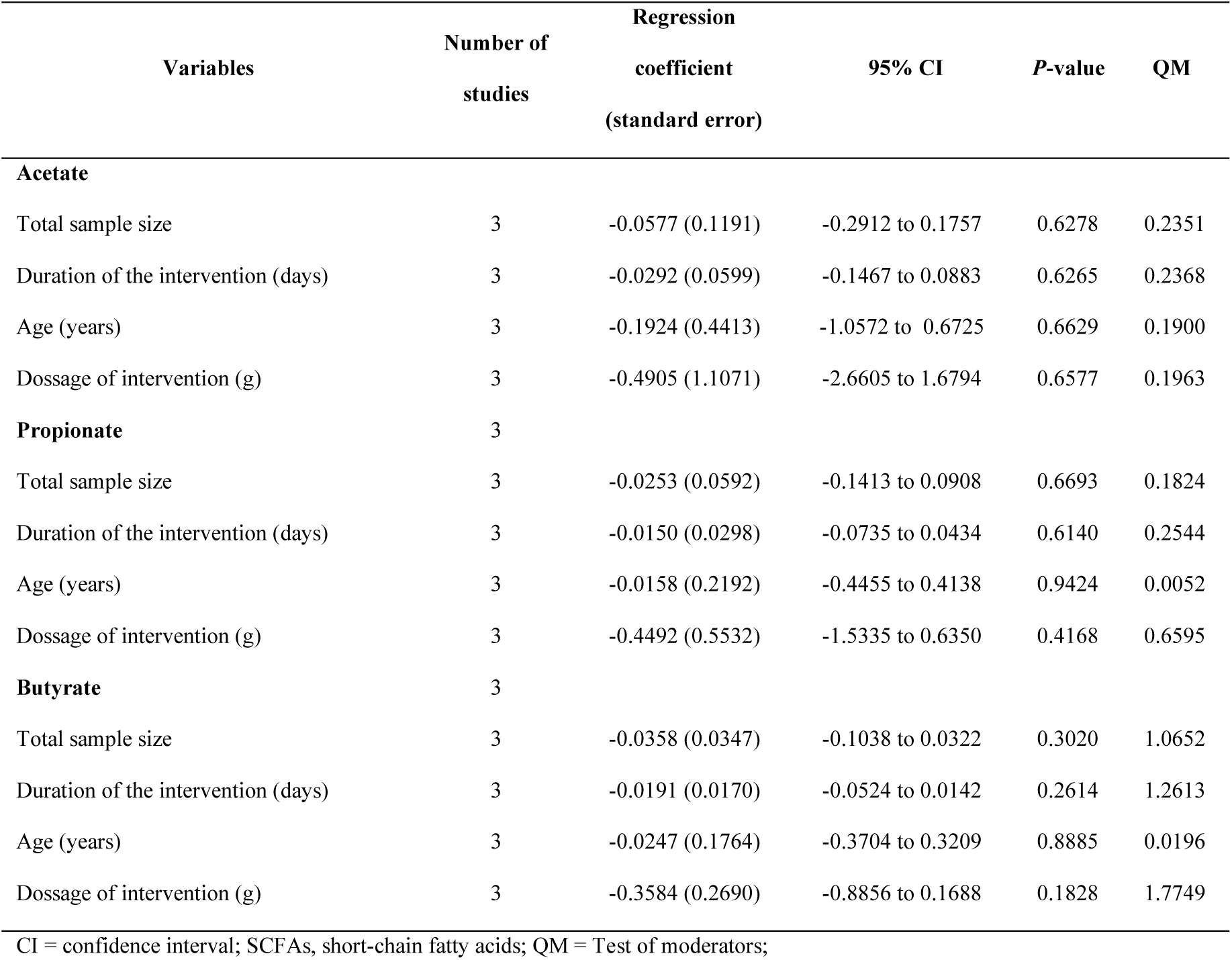
Meta-regression analysis of moderators influencing SCFAs outcomes (acetate, propionate, and butyrate) in malnourished children receiving probiotic supplementation.

#### Propionate

The use of probiotics in malnourished children may lead to a slight decrease in propionate levels compared to the control group, although the effect is not statistically significant. The meta-analysis reported a mean difference (MD) of -0.22 (95% CI: -0.76 to 0.32) based on three trials involving 126 children. Again, the confidence interval includes zero, indicating no significant difference. There was no heterogeneity among studies (I² = 0% ). Meta-regression analysis indicated that total sample size (p = 0.6693), length of the intervention (p = 0.6140), intervention dosage (p = 0.9424), and age (p = 0.4168) were not significant moderators of propionate levels, as seen in Table 2.

#### Butyrate

Probiotic therapies in malnourished children may significantly elevate butyrate levels relative to the control group. The meta-analysis reported a mean difference (MD) of 0.63 (95% CI: 0.14 to 1.12) based on three trials involving 126 children. The confidence interval does not include zero, suggesting a statistically significant increase in butyrate levels. There was no heterogeneity among studies (I² = 0% ). Meta-regression analysis indicated that total sample size (p = 0.3020), duration of the intervention (p = 0.2614), dose of intervention (p = 0.8885), and age (p = 0.1828), were not significant moderators of butyrate levels, as presented in Table 2.

#### Certainty of evidence

The certainty of evidence for the impact of probiotic supplementation on SCFAs production was assessed using the GRADE approach. Regarding SCFAs outcomes, the evidence for acetate and propionate increases was rated low due to serious risk of bias and serious imprecision. However, the evidence for butyrate increase was rated moderate due to serious risk of bias. Publication bias was undetected in all outcomes, as shown in Table 3.

**Table 3.** Summary of findings and certainty of evidence according to the GRADE approach for key outcomes.

| No of studies | Design | Risk of bias | Inconsistency | Indirectness | Imprecision | Publication bias | Experimental group | Control group | Difference (95% CI) | Final judgment |
| --- | --- | --- | --- | --- | --- | --- | --- | --- | --- | --- |
| **Acetate level** |  |  |  |  |  |  |  |  |  |  |
| 3 | RCT | Serious<sup>a</sup> | No serious | No serious | Serious<sup>b</sup> | Undetected | 63 | 63 | 0.58 (-0.51 to 1.67) | ⊕⊕⊖⊖<br>Low |
| **Propionate level** |  |  |  |  |  |  |  |  |  |  |
| 3 | RCT | Serious<sup>a</sup> | No serious | No serious | Serious<sup>b</sup> | Undetected | 63 | 63 | -0.22 (-0.76 to 0.32) | ⊕⊕⊖⊖<br>Low |
| **Butyrate level** |  |  |  |  |  |  |  |  |  |  |
| 3 | RCT | Serious<sup>a</sup> | No serious | No serious | No serious | Undetected | 63 | 63 | 0.63 (0.14 to 1.12) | ⊕⊕⊕⊖<br>Moderate |

Reasons for Downgrading the Certainty of Evidence: a, downgraded by one level for risk of bias; b, downgraded by one level due to wide confidence intervals. CI, confidence interval; GRADE, grading of recommendations assessment, development, and evaluation; RCT, randomized controlled trial.

## Discussion

The findings of this systematic review and meta-analysis confirm our hypothesis that probiotic supplemetation significantly improves SCFAs production in malnourished children. This meta-analysis indicated that therapies using probiotics may elevate acetate levels in malnourished children. Acetate is the predominant SCFA produced by enteric bacteria such as *Akkermansia muciniphila, Bifidobacterium* spp*., Ruminococcus* spp.*, Blautia hydrogenotrophica, Clostridium* spp*., Streptococcus* spp., and especially bacteria from *Bacteroidetes* phylum such as *Prevotella* spp. and *Bacteroidetes* spp., and is therefore abundantly found in both the colon and the peripheral circulation (He et al. 2021; Wu et al. 2025). Acetate is a two-carbon SCFA that can be formed through acetyl-CoA or the Wood– Ljungdahl pathway using hydrogen and carbon dioxide or formate (Mansuy-Aubert and Ravussin 2023; Wu et al. 2025). The role of acetate in overcoming malnutrition includes suppressing the inflammatory response by activating receptors. GPR43 serves as an energy provider for cells by acting as substrates for gluconeogenesis (Mansuy-Aubert and Ravussin 2023) inhibits the colonization of harmful pathogens by lowering the pH level of the gut environment (Rahayu et al. 2021) and helps maintain gut epithelial integrity by upregulating tight junction proteins, leading to reduced gut permeability (Mansuy-Aubert and Ravussin 2023). These beneficial roles of acetate may optimize the nutrition absorption in malnourished children.

This meta-analysis found that intervention with probiotic formulations had an overall positive effect on increasing propionate levels in malnourished children. Propionate is a three-carbon SCFA that can be formed through the succinate pathway, acrylate pathway, and propanediol pathway (Wenzel et al. 2018; Langfeld et al. 2021). These pathways are influenced by substrate availability, intestinal pH levels, microbial composition, intestinal gas composition, and dietary elements (e.g., types of fiber and iron content), highlighting the multifactorial nature of propionate production, which may explain the limited and inconsistent effects observed in some probiotic interventions (Martin-Gallausiaux et al. 2021). Propionate plays a multifaceted role in maintaining host metabolic and gut health (Hosseini et al. 2011), which may be particularly beneficial in improving the nutritional status of malnourished children. This SCFA has been shown to regulate the release of gut hormones such as peptide YY (PYY) and glucagon-like-peptide-1 (GLP-1) (Psichas et al. 2015). The enhancement of these hormone secretions through propionate may contribute to normalizing appetite and energy intake children (Horner and Lee 2015). Furthermore, propionate helps to modulate inflammation by activating GPR41 and GPR43 receptors and contributes to strengthening the gut barrier integrity by modulating tight junction proteins and increasing mucin production (Mansuy-Aubert and Ravussin 2023). These actions are crucial, as malnutrition is often associated with impaired gut function and increased intestinal permeability.

The increase in butyrate production following probiotic supplementation occurs through several interconnected mechanisms. Probiotics such as *Lactobacillus plantarum* Dad-13 can modulate gut microbiota composition by increasing the abundance of butyrate-producing bacteria such as *Faecalibacterium*, *Catenibacterium*, *Subdoligranulum*, and *Collinsella* (Kamil et al. 2022). These bacteria produce butyrate via specific metabolic pathways involving enzymes like butanol dehydrogenase and glutaconyl-CoA decarboxylase (Anand, Kaur, and Mande 2016). In addition, the synergistic action of probiotics and prebiotics (synbiotics) may reduce intestinal pH, thereby promoting the growth of anaerobic SCFA-producing microbes, including butyrate producers (Gunawan et al. 2021). This more acidic environment can also help suppress the growth of pH-sensitive pathogens such as *Enterobacteriaceae* (Rahayu et al. 2021).

Butyrate plays a key role in maintaining intestinal health by promoting colonocyte metabolism, enhancing mitochondrial β-oxidation, and activating PPARγ, which helps maintain an anaerobic gut environment and inhibits the growth of harmful pathogens like *Enterobacteriaceae* (Guerbette, Boudry, and Lan 2022; Hamer et al. 2008). Butyrate also stimulates GPR41 and GPR43 receptors, regulating energy metabolism, insulin sensitivity, and intestinal gluconeogenesis, all crucial for improving nutritional status (Kamil et al. 2022).

Supplementation with synbiotics, such as *L. plantarum Dad-13* and FOS, has been shown to boost SCFAs production, improve mineral absorption, and increase nutrient intake, contributing to greater weight and height in malnourished children (Gunawan et al. 2021; Gunawan et al. 2022). SCFAs, including butyrate, strengthen the intestinal barrier and modulate the immune system, playing a significant role in improving gut health and overall immunity (Rahayu et al. 2021; Rahayu et al. 2024). Therefore, elevating SCFA levels through probiotics and synbiotics has multiple positive effects on microbiota composition, energy balance, and overall nutritional improvement. Sustaining adequate butyrate levels enhances gastrointestinal health in animal models by supporting colonocyte activity, reducing inflammation, preserving gut barrier integrity, and fostering a balanced microbiome (Hodgkinson et al. 2023).

## Conclusion

This systematic review and meta-analysis demonstrates that probiotic supplementation may enhance short-chain fatty acid (SCFA) production in malnourished children, particularly butyrate. While the overall effect on total SCFAs (including acetate, propionate, and butyrate) was not statistically significant, a significant increase in butyrate levels was observed following probiotic intervention. Acetate and propionate levels also showed trends toward improvement, although these were not statistically significant, likely due to small sample sizes and methodological variability. In conclusion, probiotic supplementation shows potential as a nutritional strategy to improve SCFA production, particularly butyrate, in malnourished children. This could have important implications for restoring gut health and supporting overall growth and development. Nevertheless, further high-quality randomized controlled trials with larger sample sizes, longer follow-up periods, and standardized protocols are needed to confirm these findings and establish effective probiotic interventions for pediatric undernutrition.

## Data Availability

All data produced in the present study are available upon reasonable request to the authors

## Acknowledgment

The authors have no acknowledgment of interest to declare

## Sources of Support

The authors have no source of support of interest to declare

## Author Contributions

**Rizqi Yanuar Pauzi:** Conceptualization, Methodology, Formal analysis, Investigation, Data curation, Writing - original draft, Supervision. **Laksita Widya Kumaratih:** Conceptualization, Methodology, Investigation, Data curation, Writing - review & editing. **Puspita Tri Yuliana:** Conceptualization, Investigation, Data curation, Writing - review & editing. **Raghda Aisy Aqila:** Conceptualization, Inversigation, Data curation, Writing - review & editing.

## Author Declarations

The authors have no author declaration of interest to declare

## Notes

### Competing Interest Statement

The authors have declared no competing interest.

### Funding Statement

This study did not receive any funding

